# Using Adversarial Images to Assess the Stability of Deep Learning Models Trained on Diagnostic Images in Oncology

**DOI:** 10.1101/2021.01.17.21249704

**Authors:** Marina Z. Joel, Sachin Umrao, Enoch Chang, Rachel Choi, Daniel X. Yang, James S. Duncan, Antonio Omuro, Roy Herbst, Harlan M. Krumholz, SM Sanjay Aneja

**Affiliations:** Department of Therapeutic Radiology, Yale School of Medicine; Department of Biomedical Engineering, Yale University; Department of Neurology, Yale School of Medicine; Department of Medicine, Yale School of Medicine; Center for Outcomes Research and Evaluation at Yale (CORE)

## Abstract

**Purpose:** Deep learning (DL) models have rapidly become a popular and cost-effective tool for image classification within oncology. A major limitation of DL models is output instability, as small perturbations in input data can dramatically alter model output. The purpose of the study is to investigate the robustness of DL models in the oncologic image domain through the application of *adversarial images*: manipulated images with small pixel-level perturbations designed to assess the stability of DL models.

**Experimental Design:** We examined the impact of adversarial images on the classification accuracies of DL models trained to classify cancerous lesions across three common oncologic imaging modalities (CT, mammogram, and MRI). The CT model was trained to classify malignant lung nodules using the LIDC dataset. The mammogram model was trained to classify malignant breast lesions using the DDSM dataset. The MRI model was trained to classify brain metastases using an institutional dataset. We also explored the utility of an iterative adversarial training approach to improve the stability of DL models to small pixel-level changes.

**Results:** Oncologic images showed instability with small pixel-level changes. A pixel-level of perturbation of .004 resulted in a majority of oncologic images to be misclassified by their respective DL models (CT 25.64%, mammogram 23.93%, MRI 6.36%). Adversarial training mitigated improved the stability and robustness of DL models trained on oncologic images compared to naive models [(CT 67.72% vs 26.92%), mammogram (63.39% vs 27.68%), MRI (87.20% vs 24.32%)].

**Conclusions:** DL models naively trained on oncologic images exhibited dramatic instability to small pixel-level changes resulting in substantial decreases in accuracy. Adversarial training techniques improved the stability and robustness of DL models to such pixel-level changes. Prior to clinical implementation, adversarial training should be considered to proposed DL models to improve overall performance and safety.

## Introduction

Deep learning (DL) algorithms have the promise to improve the quality of diagnostic image interpretation within oncology (1,2). Models generated from DL algorithms have been validated across a variety of diagnostic imaging modalities including magnetic resonance imaging (MRI), computed tomography (CT), and X-ray images with classification accuracy often rivaling trained clinicians (3-9). However, the success of DL models depends, in part, on their generalizability and stability. DL algorithms, in particular, have been shown to vary output based on small changes in the input data (10,11). Such variability in response to minor changes can signal an instability in the algorithm that could lead to misclassification and problems with generalizability to different patients and settings.

Data scientists have developed strategies to quantify and mitigate the susceptibility of DL models to changes in their output in response to changes in the input using *adversarial images*. Adversarial images are manipulated images which undergo small pixel-level perturbations specifically designed to test the stability of the DL models (12-15). Pixel-level changes of adversarial images are often imperceptible to humans but can cause important differences in the model output (16-18). DL models which show stability in output when faced with adversarial images are likely the most robust and safe for clinical implementation. Previous work concerning adversarial images on DL models has largely focused on non-medical images, and the vulnerability of medical DL models is relatively unknown (18,19). Although techniques to defend against adversarial images have been proposed, the effectiveness of these methods on medical DL models is unclear.

Accordingly, we sought to test the effect of adversarial images on DL algorithms trained on three common oncologic imaging modalities. We established the performance of the DL models and then tested model output stability in response to adversarial images with different degrees of pixel-level manipulation. We then tested the utility of techniques to defend the DL models against adversarial images. This research has direct application to the use of DL image interpretation algorithms, as it provides quantitative testing of their vulnerability to small input variations and determines if there are strategies to reduce this weakness.

## Methods

### Datasets

We examined the behavior of DL algorithm outputs in response to adversarial images across three medical imaging modalities commonly used in oncology—CT, mammography, and MRI. For each imaging modality a separate DL classification model was trained to identify the presence or absence of malignancy when given a diagnostic image. Each dataset was split into a training set and a testing set in a 2:1 ratio.

CT imaging data consisted of 2,600 lung nodules from the Lung Image Database Consortium and Image Database Resource Initiative (LIDC-IDRI) collection (20). The dataset contains 1,018 thoracic CT scans collected from 15 clinical sites across the US. Lung nodules used for DL model training were identified by experienced thoracic radiologists. The presence of malignancy was based on associated pathologic reports. For patients without pathologic confirmation, malignancy was based on radiologist consensus.

Mammography imaging data consisted of 1696 lesions from the Curated Breast Imaging Subset of Digital Database for Screening Mammography (CBIS-DDSM) (21). The CBIS-DDSM contains mammograms from 1,566 patients at four sites across the US. Mammographic lesions used for DL model training were obtained based on algorithmically derived regions of interest based on clinical metadata. The presence of malignancy was based on verified pathologic reports.

MRI data consisted of brain MRIs from 831 patients from a single institution brain metastases registry (22). The presence or absence of a malignancy was identified on 4,000 brain lesions seen on MRI. Regions of interest were identified by a multi-disciplinary team of radiation oncologists, neurosurgeons, and radiologists. Presence of cancer was identified based on pathologic confirmation or clinical consensus.

To compare the relative vulnerability of DL models trained on oncologic images compared to non-medical images, two additional DL classification models were trained on established non-medical datasets. The MNIST dataset consists of 70,000 hand written numerical digits (23). The CIFAR-10 datasets includes 60,000 color images of ten non-medical objects (24).

All images were center-cropped and resized, and pixels were normalized to have unit variance. For each medical dataset, the classes (“cancer” and “noncancer”) were balanced, and data was augmented using simple data augmentations: horizontal and vertical flips as well as random rotations with angles ranging between - 20° and 20°.

### Models

For all DL classification models, we used a pre-trained convolutional neural network with the VGG16 architecture (25). Models were fine-tuned in Keras using Stochastic Gradient Descent. Details regarding model architecture and hyperparameter selection for DL model training are provided (Table S2, Table S3).

### Adversarial Image Generation

Three commonly employed first-order adversarial image generation methods—Fast Gradient Sign Method (FGSM), Basic Iterative Method (BIM), and Projected Gradient Descent (PGD)—were used to create adversarial images on the medical and non-medical image datasets (Figure 1). Each method aims to maximize the DL model’s classification error while minimizing the difference between the adversarial image and original image. All the adversarial image generation methods are bounded under a predefined perturbation size ε, which represents the maximum change to pixel values of an image. Vulnerability to adversarial images was assessed by comparing changes in model performance compared to baseline (without any adversarial images) under various perturbation sizes.

**Figure 1.**
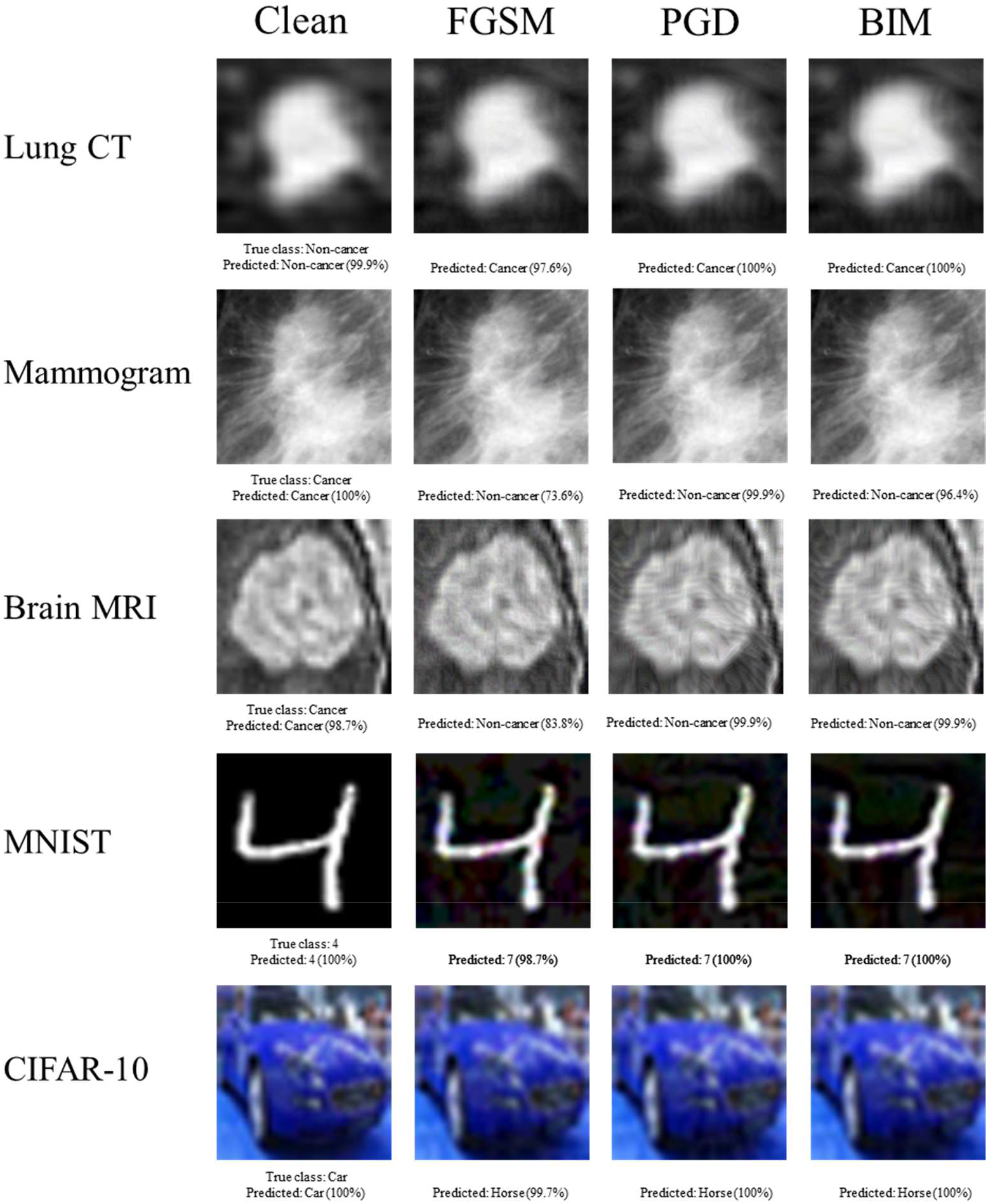
Examples of clean images and their adversarial counterparts generated using FGSM, PGD, and BIM attack methods. The percentage displayed represents the probability predicted by the model that the image is of a certain class.

The single step FGSM attack perturbs the original example by a fixed amount along the direction (sign) of the gradient of adversarial loss (15).

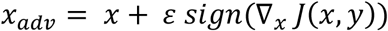

BIM iteratively perturbs the normal example with smaller step size and clips the pixel values of the updated adversarial example after each step into a permitted range (12).

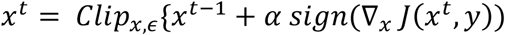

Known as the strongest first-order attack, PGD iteratively perturbs the input with smaller step size and after each iteration, the updated adversarial example is projected onto the ε-ball of x and clipped onto a permitted range (18).

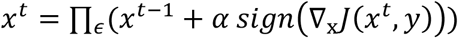

Additional information regarding adversarial image generation methods is provided (Table S1).

### Susceptibility of DL Models to Adversarial Images

We investigated the DL model performance using FGSM, PGD, and BIM adversarial image generation methods across different levels of pixel perturbation. We measured relative susceptivity to adversarial images by determining the smallest perturbation ε required for adversarial images to generate a different output. DL models which required larger pixel level perturbations are likely to be more robust and have higher levels of stability suitable for clinical implementation. Conversely, models which change outputs in response to small pixel-level perturbations are inherently unstable and potentially less generalizable across different clinical settings and patient populations.

### Adversarial Training to Improve Model Robustness

One proposed defense mechanism to prevent negative effects of adversarial images is *adversarial training*, which aims to improve model robustness by integrating adversarial samples into DL model training (18,19). By training on both adversarial and normal images, the DL model learns to classify adversarial samples with higher accuracy compared to models trained on only normal samples. We used a multi-step PGD adversarial training to increase the robustness of our DL models against adversarial attacks. In each batch, 50% of training samples were normal images, and the other 50% were adversarial images generated by PGD attack. The hyperparameters for adversarial training are detailed (Table S4). We investigated the effectiveness of our iterative adversarial training approach on the DL models trained on medical images. We measured the effectiveness of adversarial training by comparing model accuracy on adversarial samples of varying perturbation size before and after adversarial training.

### Image Level Adversarial Image Sensitivity and Model Performance

We examined each individual image’s adversarial sensitivity, as measured by the level of pixel-level perturbation necessary for DL model prediction to change as compared to an unperturbed image. We hypothesized that images requiring smaller pixel perturbations to change DL model predictions were also the images most likely to be misclassified by the model under normal conditions. By excluding images most sensitive to adversarial perturbation, we aimed to improve model performance on the remaining dataset. We identified the 20% of images most vulnerable to adversarial perturbation and excluded them from the test set. We then tested the performance of the original model on the reduced test set.

The proposed networks were implemented in Python 2.7 using TensorFlow v1.15.3 framework (26). Adversarial images were created using the Adversarial Robustness Toolbox v1.4.1 (27). The code to reproduce the analyses and results is available online at https://github.com/Aneja-Lab-Yale/Aneja-Lab-Public-Adversarial-Imaging.

## Results

### Susceptibility of DL Models to Adversarial Images

Both medical and non-medical DL models were highly susceptible to misclassification of adversarial images resulting in decreases in model accuracy (Figure 2). Medical DL models appeared substantially more vulnerable to adversarial images compared to non-medical DL algorithms. All three medical DL models required smaller pixel-level perturbations to decrease model accuracy compared to non-medical DL models (Figure 2). For example, adversarial images generated using the PGD method (perturbation = 0.002), resulted in DL model accuracy of 26.92% for CT (−48.48% from baseline), 27.68% for mammogram (−48.75% from baseline), and 24.32% for MRI (−61.81% from baseline). In contrast, adversarial images generated using the same methods/parameters did not cause substantial changes in performance for the MNIST (-.05% from baseline) or CIFAR-10 (−4.2% from baseline) trained models (Table 1). For the medical DL models, adversarial images generated using smaller pixel level perturbations (ε < 0.004) resulted in misclassification of a majority of images whereas non-medical DL models required much larger pixel perturbations (ε > 0.07 for MNIST, ε > 0.01 for CIFAR-10) for similar levels of misclassification (Table 1).

**Table 1.** Effects of adversarial attacks of varying perturbation sizes on model classification accuracy. Adversarial samples were created by FGSM, BIM, and PGD with increasing L_∞_ maximum perturbation size ε. Models for medical datasets (CT, mammogram, and MRI) required smaller attack perturbation sizes than models for non-medical datasets (MNIST, CIFAR-10) for attacks to be generally effective.

| **Attack** | **Perturbation size $\epsilon$** | **CT Accuracy (%)** | **Mammogram Accuracy (%)** | **MRI Accuracy (%)** | **MNIST Accuracy (%)** | **CIFAR-10 Accuracy (%)** |
| --- | --- | --- | --- | --- | --- | --- |
| Baseline |  | 75.41 | 76.43 | 93.64 | 99.13 | 86.13 |
| FGSM | 0.001 | 51.98 | 46.96 | 77.27 | 99.05 | 85.32 |
|  | 0.002 | 34.62 | 30.00 | 56.36 | 99.05 | 82.39 |
|  | 0.004 | 31.12 | 24.46 | 43.03 | 99.04 | 74.29 |
|  | 0.006 | 31.59 | 23.93 | 40.38 | 98.96 | 65.27 |
| PGD | 0.001 | 41.84 | 45.36 | 61.36 | 99.06 | 85.29 |
|  | 0.002 | 26.92 | 27.68 | 24.32 | 99.05 | 81.93 |
|  | 0.004 | 25.64 | 23.93 | 6.36 | 99.01 | 71.90 |
|  | 0.006 | 25.64 | 23.57 | 6.36 | 98.92 | 59.98 |
| BIM | 0.001 | 44.99 | 46.07 | 64.24 | 99.06 | 85.32 |
|  | 0.002 | 27.74 | 28.57 | 29.02 | 99.05 | 82.06 |
|  | 0.004 | 25.87 | 23.93 | 6.44 | 99.01 | 72.84 |
|  | 0.006 | 25.76 | 23.57 | 6.36 | 98.93 | 62.28 |

**Figure 2.**
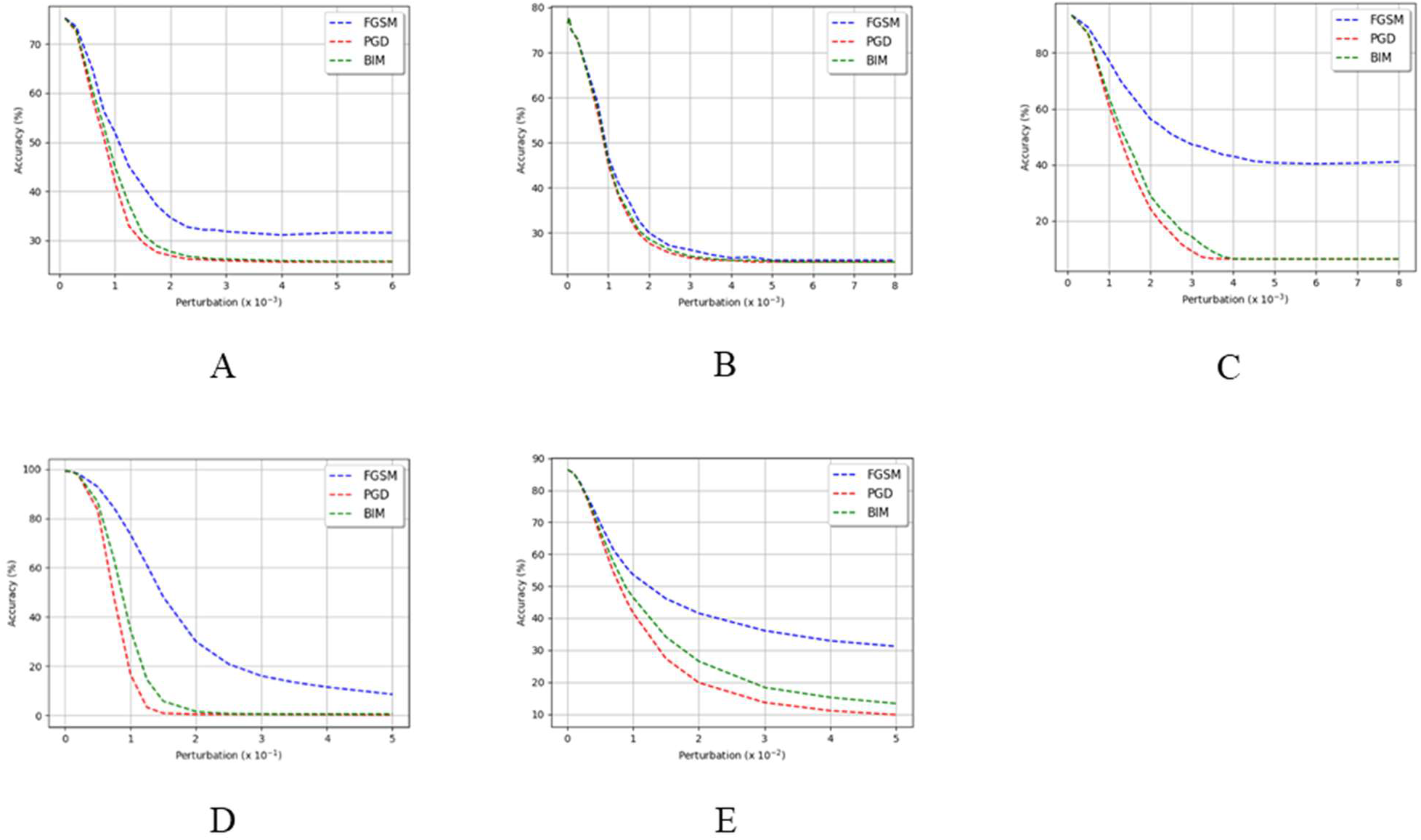
Classification accuracy of VGG16 model on adversarial examples generated by FGSM, BIM, and PGD attacks with increasing L_∞_ maximum perturbation size ε. Model performance decreased as ε increased for all datasets: A) lung CT; B) mammography; C) brain MRI; D) MNIST; and E) CIFAR-10. ^*^Note that the horizontal axis (ε) was scaled to 10^−3^ for graphs A) to C), to 10^−1^ for D), and to 10^−2^ for E).

### Adversarial Training to Improve Model Robustness

Adversarial training led to increased robustness of DL models when classifying adversarial images for both medical and non-medical images (Figure 3). Compared to baseline trained models, adversarial trained DL models caused absolute accuracy of the model on adversarial images to increase by 42.9% for CT (67.72% vs 26.92%), 35.7% for mammogram (63.39% vs 27.68%), and 73.2% for MRI (87.20% vs 24.32%) (Table S5). Despite adversarial training, DL models did not reach baseline accuracy suggesting adversarial training as only a partial solution to improve model robustness. Adversarial training became less effective when attempting to defend against adversarial images that possessed greater pixel perturbations.

**Figure 3.**
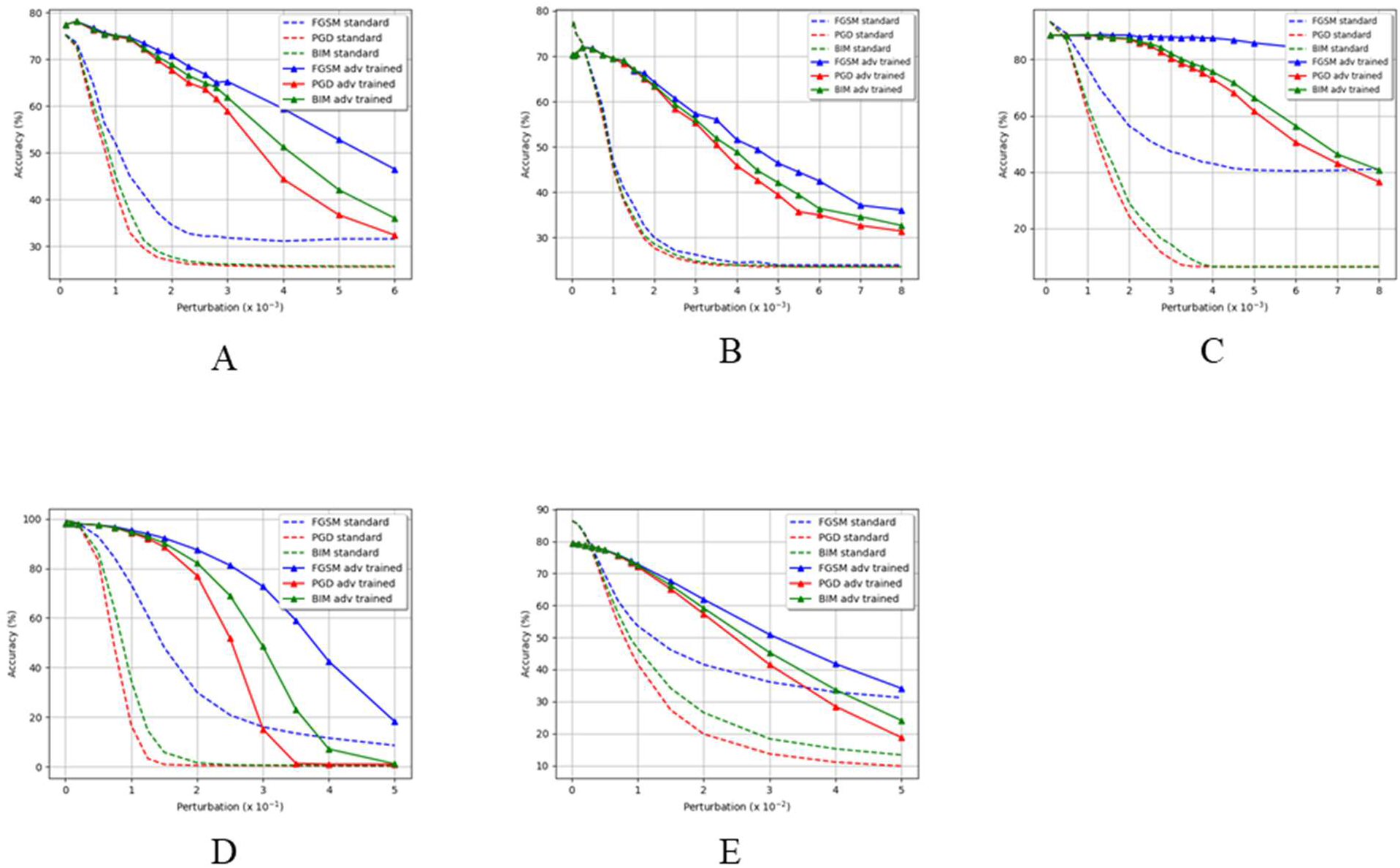
Comparison of model classification accuracy before and after adversarial training on adversarial samples crafted by FGSM, BIM, and PGD with increasing L_∞_ maximum perturbation size ε. Adversarial training significantly increased model accuracy for datasets: A) lung CT; B) mammography; C) brain MRI; D) MNIST; and E) CIFAR-10. ^*^Note that the horizontal axis (ε) was scaled to 10^−3^ for graphs A) to C), to 10^−1^ for D), and to 10^−2^ for E).

### Image Level Adversarial Image Sensitivity and Model Performance

Using image-level adversarial sensitivity, we were able to identify images most at risk for misclassification by the DL models and improve overall model performance across all diagnostic imaging modalities. Excluding the images in which the smallest pixel perturbations changed DL model outputs increased the absolute accuracy of DL models by 5.9% for CT, 3.7% for mammogram, and 5.2% for MRI (Table 2).

**Table 2.** Classification accuracy (%) of VGG16 model on the original test set and the test set excluding the 20% of test images most susceptible to adversarial attack. Images were excluded if PGD attack with perturbation size less than a certain threshold was sufficient to change the model prediction on the image. That threshold perturbation size was 0.0003 for CT, 0.00025 for mammogram, and 0.0006 for MRI.

|  | Model Accuracy (%)<br>Original Test Set | Model Accuracy (%)<br>Adversarially Aware<br>Test Set | Change in Model<br>Accuracy (%) |
| --- | --- | --- | --- |
| CT | 75.41 | 81.31 | 5.90 |
| Mammogram | 76.43 | 80.13 | 3.70 |
| MRI | 93.64 | 98.82 | 5.18 |

## Discussion

As the role of diagnostic imaging increases throughout clinical oncology, deep learning represents a cost-effective tool to supplement human decision-making and aid in image analysis tasks (28-30). However, instability of DL model outputs can limit the performance and generalizability on large-scale medical datasets and hinder clinical utility. Evaluating a proposed DL model’s susceptibility to adversarial images represents a way to identify the most robust DL models versus those at risk for erratic performance. In this study, we found that DL models trained on medical images were particularly unstable to pixel level changes from adversarial images resulting in significant decreases in expected performance. Moreover, we found diagnostic images within oncology to be more vulnerable such misclassification compared to DL models trained on non-medical images. Specifically, compared to non-medical images, all three diagnostic imaging modalities required substantially smaller pixel-level changes to reduce model performance. Furthermore, we found that adversarial training methods commonly used on non-medical imaging datasets are effective at improving DL model stability to such pixel-level changes. Finally, we showed that identifying images most susceptible to adversarial image attacks maybe helpful in improving overall robustness of DL models on medical images.

Several recent works have found that state-of-the-art DL architectures perform poorly on medical imaging analysis tasks when classifying adversarial images (14,31-35). Our work extends the findings of previous studies by evaluating performance across three common oncologic imaging modalities used for cancer detection. Additionally, we found that CT, mammography, and MRI images exhibit substantial vulnerability to adversarial images even with small pixel-level perturbations (< 0.004). We also show that DL models exhibited different levels of sensitivity to adversarial images across different imaging modalities. Furthermore, while most previous studies used only one fixed perturbation size for adversarial image attack, we varied perturbation size along a broad range to examine the relationship between model performance and attack strength.

In addition, our results corroborate previous work which showed that DL models trained on medical images are more vulnerable to misclassifying adversarial images compared to similar DL models trained on non-medical images (14,36). By using MNIST and CIFAR-10 as a control and applying the same attack settings to DL models for all datasets, we determined that DL models for medical images were much more susceptible to misclassifying adversarial images than DL models for non-medical images. One reason for this behavior could be that medical images are highly standardized and small adversarial perturbations dramatically distort their distribution in the latent feature space (37,38). Another factor could be the overparameterization of DL models for medical image analysis, as sharp loss landscapes around medical images lead to higher adversarial vulnerability (14).

In the past, adversarial training on medical DL models have shown mixed results. In some studies, adversarial training improved DL model robustness for multiple medical imaging modalities like lung CT and retinal optical coherence tomography (37,39,40). On the other hand, Hirano et al. found that adversarial training generally did not increase model robustness for classifying dermatoscopic images, optical coherence tomography images, and chest X-ray images (41). The difference in effectiveness of adversarial training can be attributed to differences in adversarial training protocols (e.g., single-step vs. iterative approaches). It’s important to note that even in studies where adversarial training showed success in improving model robustness, the results were still suboptimal, as the risk of misclassification increases with perturbation strength even after adversarial training. This is expected as adversarial training, while capable of improving model accuracy on adversarial examples, has limits in effectiveness against strong attacks even on non-medical image datasets (18).

Our work applied an iterative adversarial training approach to DL models for lung CTs, mammograms, and brain MRIs, demonstrating substantial improvement in model robustness for all imaging modalities. The effectiveness of adversarial training was highly dependent on the hyperparameters of adversarial training, especially the perturbation size for attack. While too-small perturbation sizes limit the increase in model robustness post-adversarial training, increasing the perturbation size beyond a certain threshold prevents the model from learning during training, causing poor model performance on both clean and adversarial samples. Our work demonstrated how the performance of the DL model post-adversarial training is inversely proportional to the perturbation size of the adversarial samples on which the model is evaluated. While adversarial training is effective in defending against weaker attacks with smaller perturbation magnitudes, it showed less success with attacks which altered pixels more substantially. While adversarial training proved successful at improving model performance on adversarial examples, our results were still far from satisfactory. One contributing factor is that medical images have fundamentally differently properties than non-medical images (14,37). Thus, adversarial defenses well-suited for non-medical images may not be generalizable to medical images.

We also showed that image level adversarial sensitivity, defined by the level of adversarial perturbation necessary to change image class predicted by model, is a useful metric for identifying normal images most at-risk for misclassification. This has potentially useful clinical implications as we can improve the robustness of DL models by excluding such ‘high risk’ images from DL model classification and instead providing them to a trained radiologist for examination.

There are several limitations to our study. First, we only used two-class medical imaging classification tasks. Thus, our findings might not generalize to multi-class or regression problems using medical images. Given that many medical diagnostic problems involve a small number of classes, our findings are likely still widely applicable to a large portion of medical imaging classification tasks. Our study employed only first-order adversarial image generation methods rather than higher-order methods, which have been shown to be more resistant against adversarial training (42). While the most commonly used adversarial image generation methods are first-order, there is still need for additional research on how to defend DL models for medical images against higher-order methods. A final limitation is that we used traditional supervised adversarial training to improve model robustness, while other nuanced methods like semi-supervised adversarial training and unsupervised adversarial training exist (37,43,44). While we demonstrated that supervised adversarial training is an effective method to improve model performance on adversarial examples, an interesting direction for future work would be to compare the utility of supervised adversarial training with that of semi-supervised or unsupervised adversarial training on DL models for medical images.

## Conclusion

In this work, we utilized adversarial images to explore the stability of DL models trained on three common diagnostic imaging modalities used in oncology. Our findings suggest that DL models trained on diagnostic images are vulnerable to pixel level changes which can substantially change expected performance. Specifically, we found vulnerability to adversarial images can be a useful method to identify DL models which are particularly unstable in their classifications. Additionally, we found adversarial image training may improve the stability of DL models trained on diagnostic images. Lastly, we found that image-level adversarial sensitivity is a potential way to identify image samples which may benefit from human classification rather than DL model classification. By shedding light on the stability of DL models to small pixel changes, the findings from this paper can help facilitate the development of more robust and secure medical imaging DL models which can be more safely implemented into clinical practice.

## Data Availability

All data is available from the authors upon reasonable request.

## Data Availability

Data from the study is available from the corresponding author on reasonable request.

## Supplementary Material

**Table S1.**
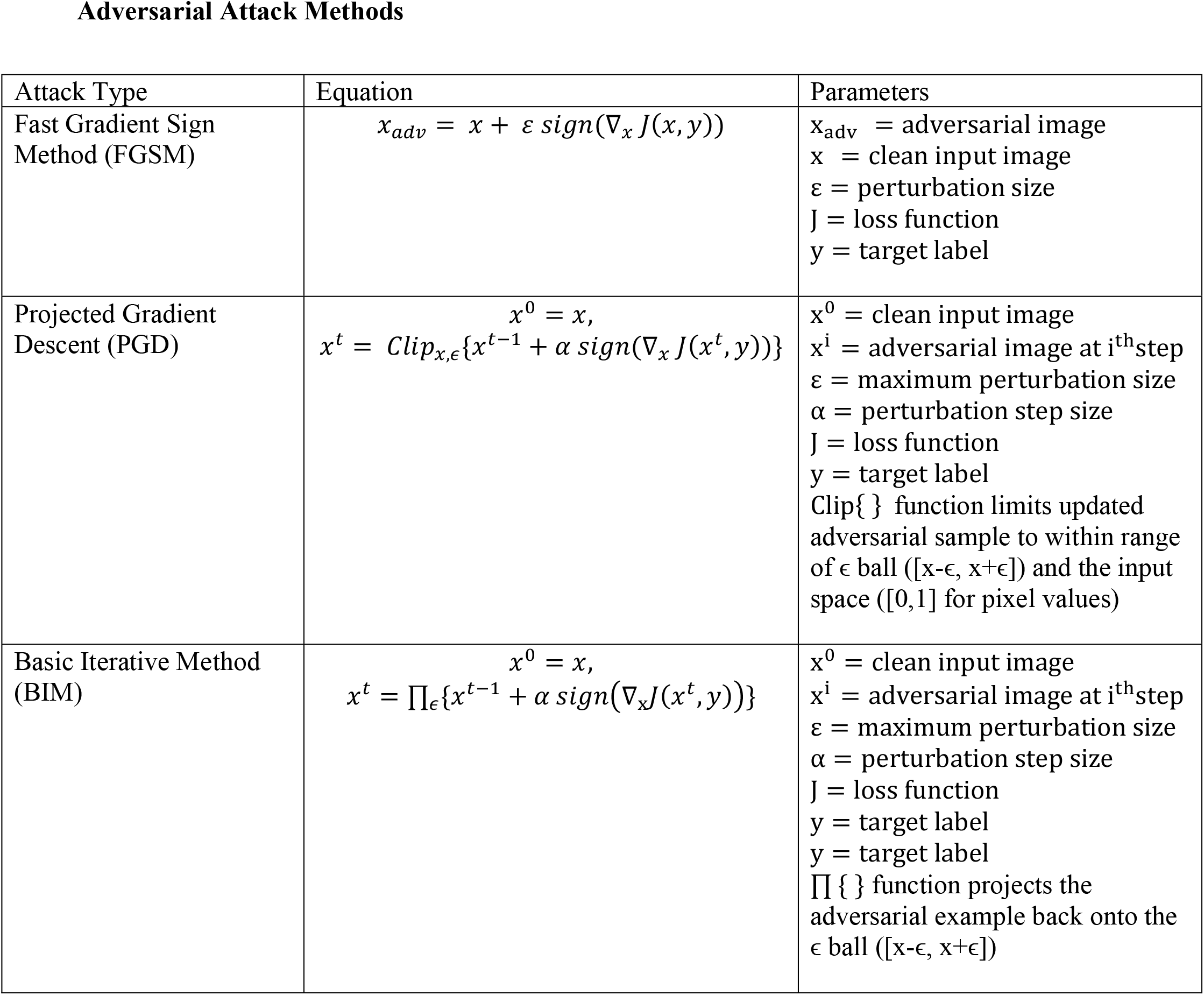
Equations and parameters for FGSM, PGD, and BIM attack methods. The number of perturbation steps for BIM and PGD are both set to 10, and the step sizes are set to ε/10 and ε/4 for BIM and PGD, respectively.

Adversarial Attack Methods
| Attack Type | Equation | Parameters |
| --- | --- | --- |
| Fast Gradient Sign Method (FGSM) | $x_{adv} = x + \epsilon \text{sign}(\nabla_x J(x, y))$ | $x_{adv}$ = adversarial image<br>$x$ = clean input image<br>$\epsilon$ = perturbation size<br>$J$ = loss function<br>$y$ = target label |
| Projected Gradient Descent (PGD) | $x^0 = x,$ $x^t = \text{Clip}_{x, \epsilon}\{x^{t-1} + \alpha \text{sign}(\nabla_x J(x^t, y))\}$ | $x^0$ = clean input image<br>$x^i$ = adversarial image at $i^{\text{th}}$ step<br>$\epsilon$ = maximum perturbation size<br>$\alpha$ = perturbation step size<br>$J$ = loss function<br>$y$ = target label<br>$\text{Clip}\{\}$ function limits updated adversarial sample to within range of $\epsilon$ ball ($[x-\epsilon, x+\epsilon]$) and the input space ($[0,1]$ for pixel values) |
| Basic Iterative Method (BIM) | $x^0 = x,$ $x^t = \Pi_{\epsilon}\{x^{t-1} + \alpha \text{sign}(\nabla_x J(x^t, y))\}$ | $x^0$ = clean input image<br>$x^i$ = adversarial image at $i^{\text{th}}$ step<br>$\epsilon$ = maximum perturbation size<br>$\alpha$ = perturbation step size<br>$J$ = loss function<br>$y$ = target label<br>$y$ = target label<br>$\Pi\{\}$ function projects the adversarial example back onto the $\epsilon$ ball ($[x-\epsilon, x+\epsilon]$) |

**Table S2.** VGG16 model architecture and parameters.

| Layer (type) | Output Shape | Parameter Number |
| --- | --- | --- |
| input_1 (Input Layer) | (None, 224, 224, 3) | 0 |
| block1_conv1 (Conv2D) | (None, 224, 224, 64) | 1792 |
| block1_conv2 (Conv2D) | (None, 224, 224, 64) | 36928 |
| block1_pool (MaxPooling2D) | (None, 112, 112, 64) | 0 |
| block2_conv1 (Conv2D) | (None, 112, 112, 128) | 73856 |
| block2_conv2 (Conv2D) | (None, 112, 112, 128) | 147584 |
| block2_pool (MaxPooling2D) | (None, 56, 56, 128) | 0 |
| block3_conv1 (Conv2D) | (None, 56, 56, 256) | 295168 |
| block3_conv2 (Conv2D) | (None, 56, 56, 256) | 590080 |
| block3_conv3 (Conv2D) | (None, 56, 56, 256) | 590080 |
| block3_pool (MaxPooling2D) | (None, 28, 28, 256) | 0 |
| block4_conv1 (Conv2D) | (None, 28, 28, 512) | 1180160 |
| block4_conv2 (Conv2D) | (None, 28, 28, 512) | 2359808 |
| block4_conv3 (Conv2D) | (None, 28, 28, 512) | 2359808 |
| block4_pool (MaxPooling2D) | (None, 14, 14, 512) | 0 |
| block5_conv1 (Conv2D) | (None, 14, 14, 512) | 2359808 |
| block5_conv2 (Conv2D) | (None, 14, 14, 512) | 2359808 |
| block5_conv3 (Conv2D) | (None, 14, 14, 512) | 2359808 |
| block5_pool (MaxPooling2D) | (None, 7, 7, 512) | 0 |
| dropout (Dropout) | (None, 7, 7, 512) | 0 |
| flatten (Flatten) | (None, 25088) | 0 |
| fc1 (Dense) | (None, 4096) | 102764544 |
| dropout_1 (Dropout) | (None, 4096) | 0 |
| fc2 (Dense) | (None, 1024) | 4195328 |
| predictions (Dense) | (None, 2) | 2050 |
| Total parameters: 121,676,610 |  |  |

**Table S3.** Training parameters for VGG16 models for all datasets.

**Model Training Parameters**
| Parameters | CNN-1<br>(CT) | CNN-2<br>(Mammogram) | CNN-3<br>(MRI) | CNN-4<br>(MNIST) | CNN-5<br>(CIFAR-10) |
| --- | --- | --- | --- | --- | --- |
| Input Image Size | 224 x 224 | 116 x 116 | 224 x 224 | 32 x 32 | 32 x 32 |
| Batch Size | 50 | 50 | 50 | 64 | 128 |
| Max Epochs | 200 | 100 | 100 | 20 | 60 |
| Patience | 50 | 20 | 5 | 0 | 10 |
| Initial Learning Rate | 0.0002 | 0.0002 | 0.0002 | 0.01 | 0.001 |
| Learning rate decay | 1e-6 | 1e-6 | 1e-6 | 0 | 1e-5 |
| Momentum | 0.9 | 0.9 | 0.9 | 0 | 0.9 |
| Loss | Binary cross-entropy | Binary cross-entropy | Binary cross-entropy | Categorical cross-entropy | Categorical cross-entropy |
| Optimizer | SGD | SGD | SGD | SGD | SGD |

**Table S4.** Adversarial training parameters for multi-step PGD training of VGG16 models.

| Parameters | CNN-1<br>(CT) | CNN-2<br>(Mammogram) | CNN-3<br>(MRI) | CNN-4<br>(MNIST) | CNN-5<br>(CIFAR-10) |
| --- | --- | --- | --- | --- | --- |
| Epsilon | 0.003 | 0.002 | 0.004 | 0.2 | 0.02 |
| Epsilon step | 0.00075 | 0.0005 | 0.001 | 0.05 | 0.005 |
| Max<br>Iterations | 7 | 7 | 7 | 7 | 7 |
| # random<br>initializations | 1 | 1 | 1 | 1 | 1 |
| Number of<br>Epochs | 200 | 100 | 200 | 50 | 100 |
| Batch size | 128 | 128 | 128 | 128 | 128 |

**Table S5.**
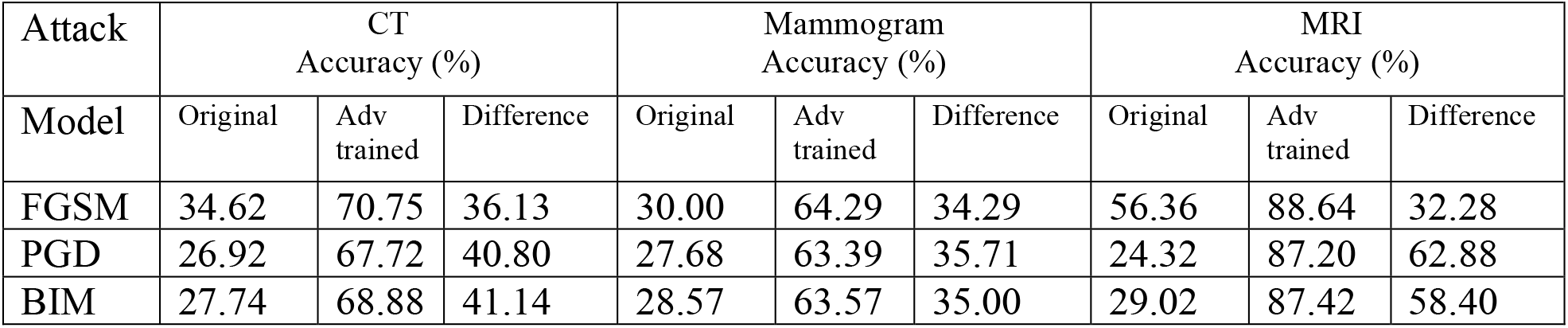
Effectiveness of Adversarial Training on Classification accuracy (%) of VGG16 model (perturbation size of .002).

| Attack | CT<br>Accuracy (%) |  |  | Mammogram<br>Accuracy (%) |  |  | MRI<br>Accuracy (%) |  |  |
| --- | --- | --- | --- | --- | --- | --- | --- | --- | --- |
| Model | Original | Adv<br>trained | Difference | Original | Adv<br>trained | Difference | Original | Adv<br>trained | Difference |
| FGSM | 34.62 | 70.75 | 36.13 | 30.00 | 64.29 | 34.29 | 56.36 | 88.64 | 32.28 |
| PGD | 26.92 | 67.72 | 40.80 | 27.68 | 63.39 | 35.71 | 24.32 | 87.20 | 62.88 |
| BIM | 27.74 | 68.88 | 41.14 | 28.57 | 63.57 | 35.00 | 29.02 | 87.42 | 58.40 |

